# Trait responsiveness to verbal suggestions predicts nocebo responding: A meta-analysis

**DOI:** 10.1101/2023.12.06.23299585

**Authors:** Madeline V. Stein, Monika Heller, Sarah Chapman, James Rubin, Devin B. Terhune

**Author notes:** Correspondence address: Madeline V. Stein Department of Psychology Institute of Psychiatry, Psychology & Neuroscience King’s College London 16 De Crespigny Park London SE5 8AB, UK. For the purposes of open access, the author has applied a Creative Commons Attribution (CC BY) license to any Accepted Author manuscript version arising from this submission.

## Abstract

Nocebo responding involves the experience of adverse health outcomes in response to contextual cues. These deleterious responses impact numerous features of mental and physical health but are characterized by pronounced heterogeneity. Suggestion is widely recognised as a contributing factor to nocebo responding but the moderating role of trait responsiveness to verbal suggestions (suggestibility) in nocebo responding remains poorly understood. We conducted a pre-registered meta-analysis (PROSPERO registration number CRD42023425605) to quantitatively synthesize available research on the relationship between suggestibility and nocebo responding. Four electronic databases were searched for original studies involving both the assessment of suggestibility and symptom reports in response to an inactive stimulus. Of 7,729 search results, 10 articles presenting 13 correlations between suggestibility and nocebo responding were analysed. A random-effects meta-analysis revealed a significant, albeit weak, positive correlation, *r*=0.21 [95% CI: 0.04, 0.37], between suggestibility and nocebo responses, such that more highly suggestible individuals displayed larger responses. Sensitivity and meta-regression analyses demonstrated that studies of higher methodological quality, including those that maintained experimenter blinding, exhibited stronger effect sizes. These results corroborate proposals that trait responsiveness to verbal suggestions confers greater response to nocebos and warrants renewed attention to the role of suggestibility in symptom induction and perception.

## Introduction

Nocebo responding is a heterogenous phenomenon involving symptom induction or exacerbation from contextual cues, such as conditioning and/or verbal suggestions, and it can occur in a diverse array of clinical, experimental, and social contexts (Colloca & Barsky, 2020; Petrovic, 2008). Nocebo responding prototypically presents as side effects or adverse events in response to medications (Heller et al., 2022), inert treatments (placebos; (Bender et al., 2023) and vaccines (Haas et al., 2022). The development of symptom expectations can occur in response to verbal or written information about potential side effects during informed consent processes (Spiegel, 1997; Wells & Kaptchuk, 2012) and patient education (Colloca & Miller, 2011; Hauser et al., 2012), and/or after observing another person displaying symptoms (Meeuwis et al., 2023). The effects of nocebo responding extend beyond the induction or exacerbation of specific symptoms. Nocebo effects can increase patient distress (Nasiri-Dehsorkhi et al., 2023), hinder treatment adherence, lead to the premature discontinuation of treatments and skew trial outcomes (Ciaramella et al., 2013; Colloca & Miller, 2011).

Responsiveness to nocebos is characterised by pronounced heterogeneity and it remains controversial whether nocebo responding is a stable trait-like variable (Kern et al., 2020; Rooney et al., 2022). Verbal suggestion, a communication for an involuntary change in experience or behaviour (e.g., “this might hurt a bit”), has been consistently demonstrated to be efficacious in the induction of nocebo responses (Petersen et al., 2014; Rooney et al., 2023; Webster et al., 2016). For example, meta-analytic evidence indicates that verbal suggestions seem to induce a nocebo response of moderate magnitude, which surpasses that of conditioning alone but falls short of the effect size produced by the joint application of both suggestion and conditioning (Rooney et al., 2023). Nocebo induction methods typically include direct verbal suggestions (e.g., “this procedure will increase your experience of pain”) or indirect verbal suggestions, which are more implicit (e.g., “some people might experience pain”) (Oakley et al., 2021; Polczyk, 2016). Trait responsiveness to direct verbal suggestions (direct verbal suggestibility) is highly stable over time (Piccione et al., 1989) and is a reliable predictor of response to hypoalgesia suggestions in both clinical and experimental contexts (Milling et al., 2021; Thompson et al., 2019). As such, the capacity to respond to verbal suggestions represents a promising candidate predictor of nocebo responding. Moreover, elevated suggestibility is a hallmark feature of a range of phenomena germane to nocebo responding, such as the experience of symptoms attributed to environmental factors (idiopathic environmental intolerance) (Stein et al., 2023), functional neurological disorder (Bell et al., 2011; Fiorio et al., 2022; Wieder et al., 2021), and mass psychogenic illness (Sapkota et al., 2020). Further indirect evidence for a link between suggestibility and nocebo responding comes from evidence showing that both trait empathy (Meeuwis et al., 2023) and absorption (Brascher et al., 2020) predict nocebo responding, and both have been found to correlate with suggestibility (Wickramasekera & Szlyk, 2003). Within the predictive processing framework (Hohwy, 2020), nocebo responses can be attributed to the formation of precise symptom priors (e.g., beliefs and expectations) that are overweighted relative to sensory information, yielding symptoms (Fiorio et al., 2022; Van den Bergh et al., 2017). By conferring greater responsiveness to suggestion, higher trait suggestibility may predispose individuals to be more likely to form strong or precise symptom priors and/or to be less likely to update these priors on the basis of sensory evidence (stubborn predictions). Although multiple studies have provided evidence of a positive link between suggestibility and nocebo responding (Corsi & Colloca, 2017; Khan et al., 2009; Winter & Braw, 2022), this effect hasn’t been replicated in others (Zech et al., 2022; Zech et al., 2020). Previous reviews and meta-analyses have largely neglected the role of suggestibility in nocebo responding and, to our knowledge, there has not yet been an attempt to formally quantify the magnitude of this relationship.

We conducted a pre-registered meta-analysis in order to quantitatively synthesize the extant literature on the association between suggestibility and nocebo responding. Toward this end, we systematically integrated research studies examining the correlation between psychometric suggestibility measures and symptom outcomes in response to an inactive (sham) procedure. We elected to omit studies with active treatments to diminish the potential confounding effect of an active treatment. In addition to quantifying the correlation between suggestibility and the magnitude of nocebo responses, we also examined whether these correlations were moderated by different measurement variables including sample type (e.g., clinical or healthy sample), nocebo suggestion type (e.g., direct or indirect verbal suggestion), target symptom domain, and various methodological features.

## Methods

We completed this meta-analysis in accordance with Meta-analyses Of Observational Studies in Epidemiology (MOOSE) (Brooke et al., 2021) and Preferred Reporting Items for Systematic Reviews and Meta-Analyses (PRISMA) guidelines (Page et al., 2021). We prospectively registered this meta-analysis on PROSPERO (registration number: CRD42023425605).

### Inclusion criteria

The data subjected to analyses met the following criteria: published in English; published in academic peer-reviewed journal; use of at least one experimental manipulation for symptom induction (e.g., verbal suggestion); use of a procedure that involved either an inactive intervention or suggestion alone that was reported as a nocebo with no intervention; and inclusion of a symptom measure. We adopted an inclusive approach when defining aversive outcomes and include outcomes from placebo studies that were not uniformly aversive (e.g., intoxication). Finally, we included papers that did not include a control condition, as we anticipated the lack of a control comparator to be a limitation of many potentially relevant studies. We had broad inclusion criteria in order to obtain an accurate representation of the experimental nocebo literature.

### Search strategy

PsycInfo, Embase, MEDLINE, and PubMed were searched in December 2022. The search was repeated in October 2023 and September 2024 and yielded no new articles to include. The search string (see Supplementary Materials) was augmented with manual searches of all included articles and relevant reviews. One author group was contacted to confirm that all relevant articles from their group had been captured by the initial search.

### Study selection

Two reviewers (MVS and MH) independently screened titles and abstracts of all retrieved articles as part of another review. One reviewer (MVS) set aside articles that met the current review’s inclusion criteria and confirmed study selection with the second reviewer (MH). Three author groups were contacted for additional data in order to be included in the analyses and were subsequently included.

### Data extraction

Data extraction was performed independently by the same two reviewers on full texts. A comprehensive list of all extracted data can be found in the supplementary methods. The primary outcomes were correlation coefficient(s) between a suggestibility scale and nocebo response (operationalised as nocebo condition alone [no control], nocebo-control, or nocebo-baseline). When multiple correlations from a single suggestibility scale with subscales were reported, we only extracted the correlation for the total scale score. Additionally, studies were coded for the following criteria: suggestibility scale administration context (i.e. hypnosis-related context or not); suggestibility scale administration type (live vs. recording); scale type (direct, indirect, or not applicable); scale administration context (group vs. individual); and good psychometric properties for the suggestibility scale (yes vs. no), based on relevant reliability and validity data presented in the paper. The reviewers displayed good agreement (86%); any discrepancies were resolved through discussion with DBT.

### Study methodological quality

A 15-item scale was developed to assess study methodological quality (see Supplementary Methods). Items were adapted from a previous measure (Thompson et al., 2019; Wieder et al., 2021) and included items based on Cochrane criteria and PRISMA recommendations. The two reviewers independently rated each item categorically (0=criterion not met, 1=criterion met), with any discrepancies resolved through discussion with DBT, and a total score (percentage of criteria met) was computed for each study. Agreement between reviewers was good (percentage agreement: 82%; Cohen’s kappa=.62).

### Data synthesis

All analyses were performed using Jamovi (The jamovi project, 2023). Individual study effect sizes included unadjusted correlation coefficients between suggestibility scale scores and nocebo responses that were transformed to *z*-scores using Fisher’s *r*-to-*z* transformation (*Z_r_*) and analysed using random-effects meta-analysis with the DerSimonian-Laird method. Alongside aggregate *Z_r_* values, we report raw correlation coefficients for ease of interpretation. Heterogeneity was computed through I^2^ and τ^2^, where I^2^ >50% indicates moderate or greater heterogeneity and τ describes the variance of true effect sizes, with values >10% indicating greater heterogeneity. We assessed publication bias by examining funnel plots of effect sizes against standard errors for asymmetry and tested for asymmetry using Egger’s bias test (Egger et al., 1997), where *p*<.10 is suggestive of asymmetry. We also estimated asymmetry-corrected cumulative effect sizes using the trim-and-fill method (Duval & Tweedie, 2000).

In order to account for heterogeneity in effect sizes, we performed subgroup analyses and a series of meta-regression analyses using binary and continuous moderators when there was a minimum of two studies per moderator level and ten studies total, respectively. Moderator variables with >2 levels were decomposed into simpler 2-level moderators. Moderators included 18 categorical variables (0=absent/no, 1=present/yes unless otherwise specified): study design (0=between-groups 1=design within-subjects), blinding of participant and/or experimenter to experimental condition, nocebo suggestion target symptom domain (pain, dyspnoea, dizziness, nausea, motor inhibition, cognitive function; each symptom coded as 0=absent vs. 1=present, e.g. no pain vs. pain), use of conditioning procedure, suggestion administration type (verbal, video, non-verbal), suggestibility scale type (0=indirect, 1=direct), nocebo administration procedure (pill, cream, procedure, nasal spray, inhalation, injection; each administration procedure coded as 0=absent vs. 1=present, e.g. no pill vs. pill), control comparator (control condition, baseline, or none), presence of hypnotic context, scale administration context (0= individual, 1=group); and one continuous variable: methodological quality percentage score.

## Results

### Study inclusion

A PRISMA diagram presenting study selection can be found in Fig. 1. The final sample of 10 included papers reported 13 suggestibility-nocebo response correlation pairs with three papers reporting two correlations. Details of the included studies can be found in Table 1 and a full list of included papers can be found in the Supplementary Results.

**Figure 1.**
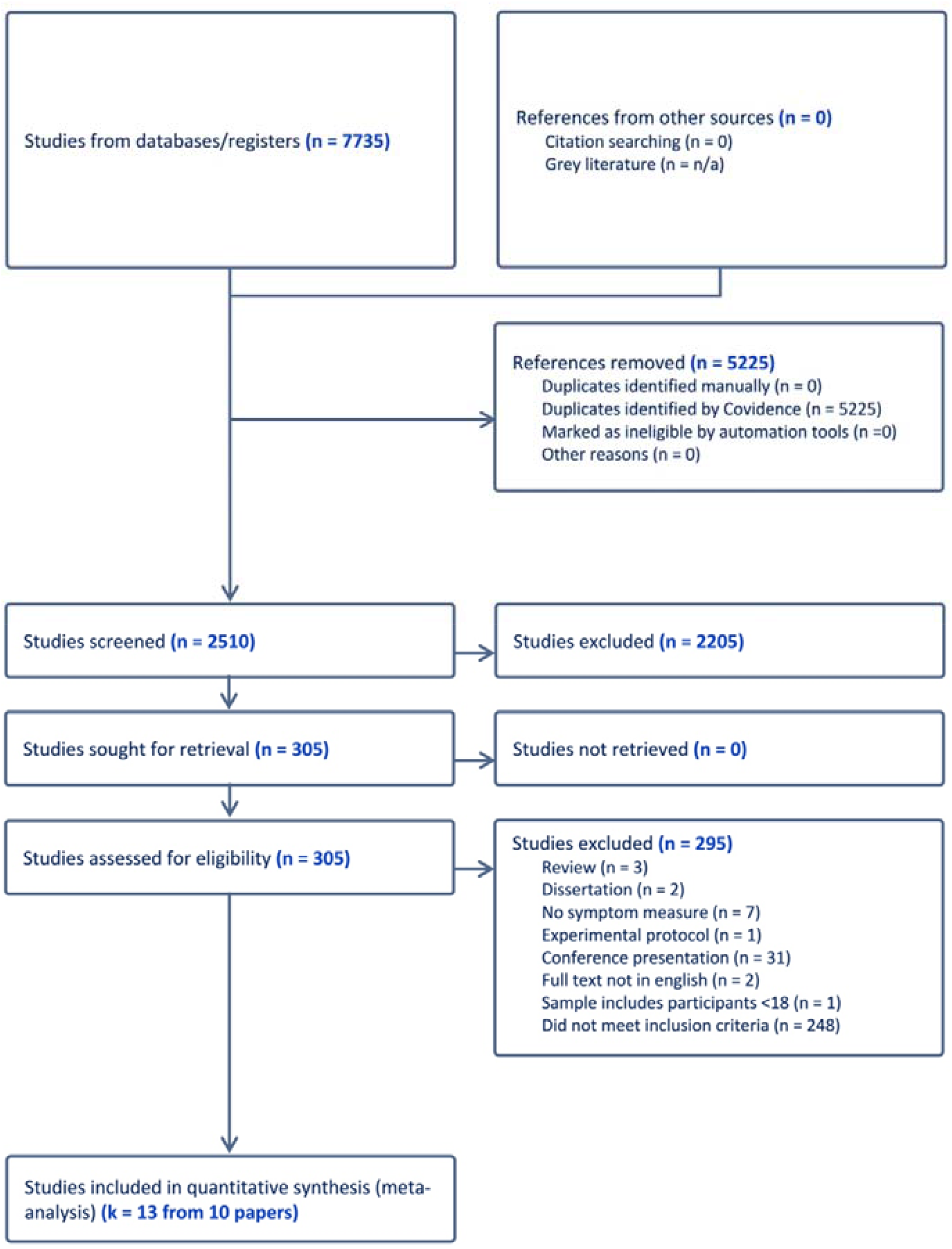
*PRISMA flowchart of the study selection process*.

**Table 1.** Principal features of included papers reporting associations between suggestibility and nocebo responding (n=10).

| Source | Country | Study design | Sample | N (% female) | Age M (SD) | Symptom domain | Symptom outcome measure | Experimental condition | Control condition | Suggestibility scale |
| --- | --- | --- | --- | --- | --- | --- | --- | --- | --- | --- |
| Corsi & Colloca, 2017 | US | Within | Healthy volunteers | 46 (52%) | 27.41 (1.07) | Pain | VAS pain | Verbal suggestion; conditioning procedure | Condition: conditioned control cue | MISS |
| Di Stefano et al., 2022 | - | Between | GI patients | 40 (87%) | 39 (16) | Pain, dyspnoea, nausea | VAS GI symptom scale | Verbal suggestion; sham lactose test | Condition; sham lactose test | MISS |
| Khan et al., 2009 | - | Between | Patients experiencing seizures | 47 (58%) | - | Seizure | Symptom induction | Verbal suggestion | - | HIP |
| Leigh et al., 2003 | CA | Within | Asthma patients | 17 (64%) | 46.8 (14.25) | Dyspnoea | Borg Dyspnoea Scale | Verbal suggestion; saline inhalation | Baseline | CIS |
| Sharav et al., 2023 | - | Mixed | Healthy volunteers | 24 (62%) | 26.65 (-) | Pain | VAS pain | Verbal suggestion; conditioning | - | SHALIT |
| Winter & Braw, 2021 | IL | Between | Recovered COVID-19 patients | 90 (75%) | Nocebo: 38.60 (1.61) Control: 39.84 (1.76) | Cognitive performance | Cognitive Failures Questionnaire | Textual suggestion | Control group; textual suggestion | SSS |
| Woody et al., 1997* | CA | Between | Undergraduate students | 93 (44%) | 20.5 (-) | Dizziness, nausea, motor inhibition, cognitive function | Subjective experience measure; Unsuggested Symptom Checklist | Verbal suggestion; sham alcohol | - | HGSHS:A |
| Zech et al., 2019* | DE | Within | Healthy, non-medical professionals | 46 (54%) | 34.3 (15.2) | Motor performance | Maximum arm muscle strength (% decrease from baseline) | Verbal suggestion | Baseline | HGSHS:A |
| Zech et al., 2020* | DE | Within | Elective surgery patients | 45 (55%) | 43.8 (15) | Motor performance | Maximum arm muscle strength (% decrease from baseline) | Verbal suggestion | Baseline | HGSHS-5 |
| Zech et al., 2022 | DE | Mixed | Healthy volunteers | 50 (58%) | 29.1 (12.7) | Motor performance | Spirometry | Verbal suggestion | Baseline | HGSHS-5 |

Notes: \* = reports 2 correlation coefficients with no variation in principle features; - = Not reported; Within = within-groups design; Between = between-groups design; Mixed = mixed experimental design; US= United States; CA= Canada; IL= Israel; DE=Germany; CIS = *Creative Imagination Scale*; GI = Gastro-Intestinal; HGSHS:A = *Harvard Group Scale of Hypnotic Susceptibility: Form A*; HGSHS-5 = *Harvard Group Scale of Hypnotic Susceptibility-5 item*; HIP = *Hypnotic Induction Profile*; MISS = *Multidimensional Iowa Suggestibility Scale*; SHALIT = *Six-minute Arm Levitation Test*; SSS = *Short Suggestibility Scale*; VAS = *Visual Analogue Scale*

### Methodological quality criteria

Methodological quality scores varied across the sample of included papers (*n*=10), with most tending to meet around half of the criteria (M%±SD: 61±13; range: 40-80; see Supplementary Table 1). There was no variability in methodological scores and ratings across studies extracted from a single paper. All studies used a reliable and valid suggestibility scale, and the majority (90%; *n*=9) included a referenced symptom measure. However, a little more than half of the included papers (60%; *n*=6) ensured appropriate experimenter blinding to participants’ suggestibility level or nocebo response, whereas 50% (*n*=5) of the papers ensured participants were adequately blinded.

### Meta-analysis of bivariate correlations between nocebo response and suggestibility

A random-effects meta-analysis of correlation coefficients (Fisher’s *r*-to-Z [*Z_r_*]) between symptom outcomes in response to an inactive (sham) procedure and suggestibility scale scores (*k*=13; see Figure 2) revealed a significant association, *Z_r_*=0.22 [95% CI: 0.04, 0.38], Z=2.48, *p=*.013, corresponding to a weak correlation, *r*=.21 [0.04, 0.37]. As can be seen in Figure 2, there was considerably heterogeneity in effect sizes, I^2^=78%, τ^2^=.07, with correlations ranging from *Z_r_*=-0.32 to *Z_r_*=0.73 (*r* range: -0.31 to 0.62). A Jackknife analysis, in which each correlation pair was sequentially omitted and the analysis was reperformed, confirmed the aggregate effect size varied in the weak range (*Z_r_* range: 0.13-0.26). The cumulative effect remaining statistically significant in all of the 13 iterations, indicating that it was not driven by a single study. These results demonstrate that suggestibility is a significant, reliable, predictor of nocebo responding.

**Figure 2.**
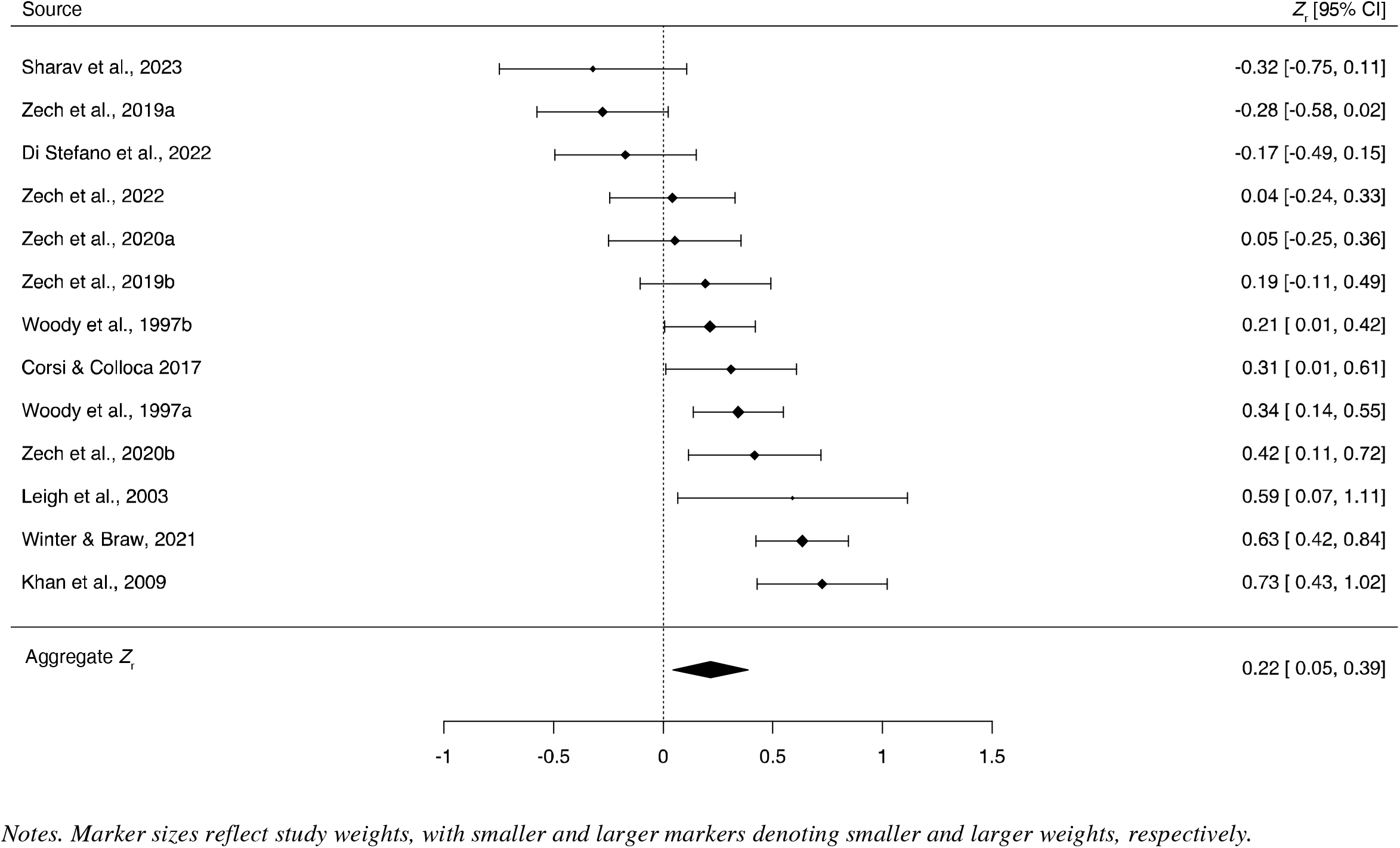
Forest plot of correlation coefficients (*Z_r_*) *[95% CIs] between suggestibility and nocebo responses (k=13)*.

Given the relatively low average methodological quality score (∼60%), and the high level of heterogeneity in effect sizes, we next performed a series of sensitivity analyses to determine whether the aggregate effect size would be stable in more methodologically robust studies. Toward this end, we repeated the central analyses on subgroups of studies with superior methodological features; we only conducted these subgroup analyses when three or more studies could be included, resulting in four analyses. First, we restricted the analysis to studies with higher methodological quality scores (rating>60%; *k*=7), the majority of the studies included in this subgroup (*k*=6) were also well-powered to detect moderate-to-large effect sizes (*n*≥46). This resulted in a larger overall effect size, *Z_r_*=0.41 [0.23, 0.59], Z=4.54, *p*<.001, equivalent to *r*=.39, which fell outside the 95% CIs of the original correlation, with a modest reduction in heterogeneity, I^2^=67% and τ^2^=.03. Similarly, studies that implemented participant condition blinding and experimenter blinding of condition, suggestibility level, and nocebo response (*k*=7), demonstrated a larger, and highly significant, effect size, *Z_r_*=0.37 [0.16, 0.58], Z=3.46, *p<*.001, equivalent to *r*=.35, with only a minor decrease in heterogeneity, I^2^=76% and τ^2^=.05. By contrast, studies that used a measure of direct verbal suggestibility (*k*=10) demonstrated a comparable, significant effect to the full sample, *Z_r_*=0.20 [0.01, 0.38], Z=2.09, *p=*.036, equivalent to *r*=.19, with a minor decrease in heterogeneity, I^2^=74% and τ^2^=.06. In a similar manner, studies that included a control condition (*k*=9) exhibited a weak effect size that was comparable to that of the total sample, albeit non-significant, *Z_r_*=0.19 [-0.03, 0.41], Z=1.72, *p*=.085, equivalent to *r*=.18, with no reduction in heterogeneity, I^2^=78% and τ^2^=.08. Similarly, studies that used behavioural/physiological outcomes (*k*=6), yielded an effect size comparable to the overall main effect, *Z_r_*=0.19 [-0.08, 0.46], Z=1.36, *p*=.17, with moderate heterogeneity, I^2^=80%, τ^2^ =.09.

Cumulatively, these analyses indicate that the utility of suggestibility as a predictor of nocebo responding is relatively stable when the data are restricted to methodologically superior studies, and indeed is substantially greater among the most methodologically rigorous studies.

### Publication bias

Egger’s test was used to assess publication bias and did not reveal significant funnel plot asymmetry, Z=-0.73, *p*=.46, in the total sample (Figure 3), suggesting that there was no clear evidence for publication bias. No values were calculated using the trim-and-fill method; therefore, none were applied.

**Figure 3.**
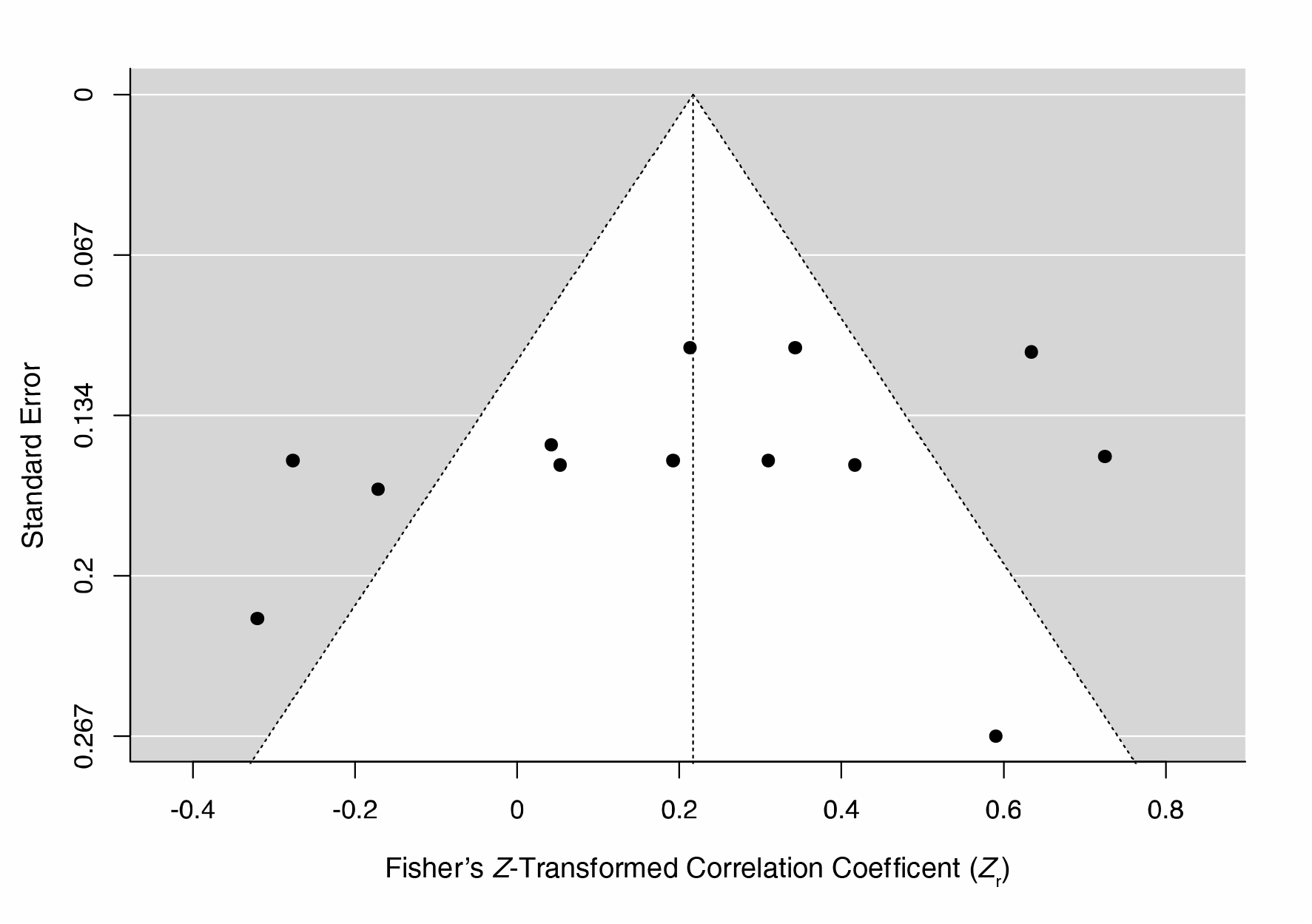
Funnel plot of nocebo-suggestibility correlation coefficients (Z_r_) as a function of standard error (k=13). Markers denote individual study effect sizes.

### Meta-regression analyses

Owing to the moderate heterogeneity in effect sizes in the total sample and subgroups, we undertook a series of meta-regression analyses to identify the variables that moderated the magnitude of these associations (see Table 2 and Supplementary Table 3). We first considered the potential moderating effects of individual methodological quality criteria. Due to our inclusion criterion of at least two studies per moderator level (e.g., unblinded v. blinded experimenter), seven individual methodological quality items were eligible for analysis and were included as binary moderators of correlation coefficients (Table 2). Among these criteria, the only significant moderator of effect sizes was experimenter blinding to the suggestibility level or nocebo response of the participants. In particular, experimenter blinding was a highly significant, positive moderator, such that studies that ensured that the experimenter was blinded exhibited larger correlations between suggestibility and nocebo responding. Consistent with this result, the overall methodological quality score was also a highly significant continuous moderator; this effect was also positive in direction, although the effect size was weak in magnitude. Both results align with the preceding results of our sensitivity analyses. Taken together these results suggest that studies that were of higher methodological quality, in particular those that implemented experimenter blinding, tended to display a higher correlation between nocebo responding and suggestibility.

**Table 2.** Meta-regression analyses on correlation coefficients between suggestibility and nocebo responding (k=13).

| Moderator | ($\Delta Z_r$) [95% CIs] | Z | p | I<sup>2</sup> |
| --- | --- | --- | --- | --- |
| Item 1: Reliability data presented in paper | 0.22 [-0.17, 0.61] | 1.08 | .28 | 77% |
| Item 3: Clear sample recruitment site | 0.23 [-0.18, 0.65] | 1.11 | .26 | 78% |
| Item 4: Clear participant recruitment | 0.16 [-0.31, 0.65] | 0.68 | .49 | 79% |
| Item 8: Clear protocol reporting | 0.31 [-0.15, 0.76] | 1.33 | .18 | 76% |
| Item 9: Experimenter blinding to suggestibility or nocebo response | 0.45 [0.18, 0.71] | 3.35 | <.001 | 61% |
| Item 10: Participant blinding | 0.19 [-0.07, 0.47] | 1.44 | .15 | 78% |
| Item 11: Experimenter condition blinding | 0.14 [-0.38, 0.66] | 0.53 | .59 | 78% |
| Total methodological quality score | 0.02 [0.01, 0.03] | 3.36 | <.001 | 61% |
| Pain as symptom outcome | -0.32 [-0.73, 0.07] | -1.58 | .11 | 77% |
| Use of conditioning | -0.22 [-0.72, 0.28] | -0.85 | .39 | 79% |
| Use of control comparator | -0.07 [-0.46, 0.31] | -0.38 | .69 | 79% |
| Use of direct symptom suggestion | 0.04 [-0.49, 0.41] | -0.19 | .85 | 80% |

Notes: $\Delta Z_r$ = Fisher's *r*-to-*z* correlation difference; all moderators were binary except Total methodological quality score; item refers to the methodological quality criteria (see Supplementary results); for all binary moderators, 0=variable absent, 1=variable present.

We next examined whether 16 binary variables moderated correlation coefficients in the total sample (Table 2 and Supplementary Table 3). There were no significant moderating effects in relation to target symptom domain, suggestibility measurement context, such as within a hypnotic context, suggestion type or suggestibility scale type. Due to the limited sample size these non-significant results should be interpreted cautiously; however, they suggest that the association between suggestibility and nocebo responding is not significantly contingent upon these variables.

Considering the largely unaccounted for heterogeneity, and diverse selection of suggestibility measures, we undertook a series of exploratory meta-regression and subgroup analyses to better understand how the choice of suggestibility scale might account for the observed variability. In particular, we distinguished three types of suggestibility scales: established measures of direct verbal suggestibility (*k*=5), *Z_r_*=0.35 [0.13, 0.57], Z=3.12, *p*=.002 [*r*=.38], I^2^=68%, τ^2^=.04, abbreviated measures of direct verbal suggestibility (*k*=5), *Z_r_*=0.03 [-0.24, 0.29], Z=0.17, *p*=.86 [*r*=.02], I^2^=71%, τ =.07, and retrospective self-report measures of suggestibility (*k*=3), *Z_r_*=0.27 [-0.19, 0.73], Z=1.14, *p*=.24 [*r*=.25], I^2^=89%, τ^2^=.11. Meta-regression analyses indicated that nocebo correlations with the established scales displayed a non-significant trend towards being greater than those with the abbreviated scales, Δ*Z_r_*=0.33 [-0.01, 0.67], Z=1.86, *p*=.062 [Δ*r*=.32], I^2^=70%, τ^2^=.05. By contrast, correlations with the established direct scales did not significantly differ from those with retrospective scales, Δ*Z_r_*=0.08 [-0.36, 0.53], Z=0.36, *p*=.71 [Δ*r*=.08], I^2^=83%, τ^2^=.06, nor did they differ between abbreviated direct scales and retrospective scales, Δ*Z_r_*=0.25 [-0.25, 0.75], Z=0.98, *p*=.33 [Δ*r*=.23], I^2^=80%, τ^2^=.09. Taken together, these results imply that the observed association between nocebo responding and suggestibility was primarily driven by established direct verbal suggestibility scales and to a lesser extent retrospective self-report scales.

## Discussion

Our meta-analysis quantified the relationship between suggestibility and nocebo responding by synthesizing data from available empirical studies. The analyses demonstrated that suggestibility is a reliably significant, albeit weak, positive predictor of nocebo responding. Notably, the magnitude of this effect was greater in studies with superior methodological rigour (and larger sample sizes), particularly those that implemented experimenter blinding. These data complement previous systematic reviews and meta-analyses showing the clear efficacy of verbal suggestion in the induction of nocebo responses (Petersen et al., 2014; Rooney et al., 2023; Webster et al., 2016) but bridge a gap in the literature by illustrating the significant moderating influence of trait responsiveness to suggestion on nocebo responding. This aligns with a wealth of data showing that trait suggestibility is a reliable predictor of responses to suggestion-based interventions (Jensen et al., 2023; Milling et al., 2021; Thompson et al., 2019) but also seems to function as a risk factor for conditions and symptoms adjacent to nocebo responding including functional neurological disorder (Wieder et al., 2021), mass psychogenic illness (Sapkota et al., 2020) and symptoms associated with environmental factors (idiopathic environmental intolerance) (Stein et al., 2023).

The frequently overlooked role of suggestibility in predicting nocebo responding has multiple salient implications for the empirical study of nocebo responding, as well as the occurrence of nocebo effects in clinical practice. Despite the cumulative observed association between suggestibility and nocebo responding, this effect was characterised by moderate heterogeneity that remained mostly unaccounted for in our sensitivity and meta-regression analyses. This unaccounted-for heterogeneity is potentially partly attributable to undetected publication bias. Although publication bias as a source of the observed effect cannot be ruled out, multiple studies included here reported non-significant correlations and suggestibility within our sample was most often a secondary point of interest, thereby minimising the motivation to avoid reporting non-significant suggestibility-nocebo correlations. The inter-study variability is plausibly attributed to methodological differences across studies, the influence of which was not observed because of the overall small sample size. Some plausible moderators of this association include the characteristics of nocebo suggestions, suggestibility instruments, and target symptom domain.

Although the efficacy of verbal suggestion in nocebo responding is well established (Petersen et al., 2014; Rooney et al., 2023; Webster et al., 2016), the nuances of suggestion have received only scant attention in research on nocebo effects. For example, verbal suggestions (“your pain will reduce”) and instructions (“try to reduce your pain”) are frequently conflated (e.g., Rooney et al., 2023), even though they are hypothesized to reflect distinct phenomena (Brass et al., 2017; Terhune et al., 2017). Similarly, researchers have tended to neglect important differences in the phrasing of verbal suggestions, such as the distinction between direct and indirect suggestions. Using meta-regression analyses, we aimed to disentangle the role of suggestion type, though our results should be considered preliminary as we found no significant evidence for moderating effects. In our sample of included studies, the specific verbal suggestions used to induce nocebo effects were only rarely reported, which hindered our ability to reliably code suggestion type. The moderating influence of suggestibility is plausibly contingent on the suggestion type administered. Accordingly, we recommend future studies report the exact wording of suggestions and provide specific details about when suggestions were administered during an experiment. This approach will enhance the reproducibility of experimental research in this domain and permit future analyses on the moderating influence of different features of verbal, textual, and non-verbal suggestions (see also Halligan & Oakley, 2014).

Another plausible source of heterogeneity in the association between suggestibility and nocebo responding is variability in the psychometric measurement of suggestibility. The domain of suggestibility includes responsiveness to direct and indirect verbal suggestions, often measured through work-sample instruments (e.g., Woody & Barnier, 2008). Little consideration has been devoted to salient differences between suggestibility measures, with most researchers implicitly assuming a uniform construct of suggestibility. In nocebo studies, trait suggestibility has been measured with established work-sample instruments or retrospective self-report measures. The former (direct verbal suggestibility scales) typically draw upon measures from experimental hypnosis research, in which responsiveness to a set of verbal suggestions for changes in motor control, cognition and perception are administered by an experimenter and then evaluated with brief tests (Acunzo & Terhune, 2021; Woody & Barnier, 2008). In our sample, 10 studies used direct verbal suggestibility scales, however, there was considerable variability in the number and type of suggestions included. Of note, the studies (Khan et al., 2009; Leigh et al., 2003; Woody et al., 1997; Zech et al., 2019) that used established, longer direct verbal suggestibility scales (Barber & Wilson, 1978; Shor & Orne, 1963) yielded non-significantly larger correlations than the studies (Sharav et al., 2023; Zech et al., 2022; Zech et al., 2020) that used abbreviated direct verbal suggestibility scales (Hilgard et al., 1979; Riegel et al., 2021). It remains unclear whether these differences are due to a greater scoring range in the established scales, or their inclusion of cognitive-perceptual suggestions, which are plausibly more relevant to symptom induction than the mostly motor suggestions on the abbreviated scales. On the other hand, the use of retrospective self-report measures of suggestibility (Kotov et al., 2004) yielded more ambiguous results. Although preliminary, these results suggest that future research will benefit from the inclusion of more established suggestibility scales that include a more diverse set of suggestions and the avoidance of abbreviated suggestibility scales.

Nocebo symptoms can be reliably induced but their severity is inconsistent across symptom domains (Bagaric et al., 2022; Meeuwis et al., 2023; Rooney et al., 2023; Wolters et al., 2019). We assessed several symptom domain moderators: pain, dyspnoea, dizziness, nausea, motor inhibition, and cognitive function, all of which were found to be non-significant, which indicates that the nocebo- suggestibility association was not greater in these domains than in all others. This is likely due to the relatively small sample of included studies, such that the moderation analyses plausibly lacked sufficient power to detect weak correlation differences, especially given that each symptom domain included only 2-4 studies. Alternatively, it is plausible that the diversity in nocebo induction methods could account for the lack of moderation effects. For example, suggestibility was previously found to correlate with placebo responding when suggestion alone was used to induce placebo hypoalgesia (Parsons et al., 2021). As such, it might be that the role of suggestibility in predicting nocebo responding is dependent on suggestion-centred symptom induction techniques. Future research would benefit from clarifying how suggestibility relates to nocebo effects from different nocebo induction methods and corroborate the results of previous reviews by disentangling the moderating effects of target symptom domain within nocebo responding.

The results of this meta-analysis complement previous research demonstrating links between suggestibility and phenomena germane to nocebo responding and corroborate proposals that suggestibility confers risk for somatic symptom reporting and nocebo responding more broadly (Corsi & Colloca, 2017; Spiegel, 1997). As such, highly suggestible individuals may be more susceptible to nocebo responses due to potential side effect information, informed consent processes, or treatment misinformation. From a predictive processing perspective (Hohwy, 2020), highly suggestible individuals may have a capacity for forming more precise symptom priors that shape perception in the direction of contextual cues (Fiorio et al., 2022; Van den Bergh et al., 2017). For example, during treatment information processes, side effect information may function as a type of indirect verbal suggestion leading to the formation of precise symptom priors, inducing premature or imitative adverse events (i.e., nocebo response). These symptom priors might compound and generalize, from treatment-specific nocebo responding, to more pronounced functional conditions depending on other individual difference factors (e.g., treatment history). Future research should explore the interplay between suggestibility, contextual cues, and the formation of precise symptom priors, investigating how individual differences and life experiences might modulate the transition from treatment-specific nocebo responses to broader functional conditions. Additionally, future research should examine suggestibility as a predictor of response to active treatments (Nitzan et al., 2015; Szigeti et al., 2024)

### Limitations

The presented results should be interpreted within the context of their limitations. As in the broader experimental study of nocebo effects, the majority our sample relied on self-report measures of symptom outcomes (*k*=7), which are susceptible to reporting biases. However, we partially accounted for this possibility in our sensitivity analyses which suggest that the overall main effect is unlikely to be driven by the use of self-report measures. Although we observed stronger effects in higher-quality studies, the overall methodological quality across studies was relatively low with no study scoring >80%. In particular, very few of the studies included control conditions and many used baseline as a control comparator. Future research examining the role of suggestibility in nocebo responding will need to employ more rigorous designs and ensure clear reporting of experimental parameters including the precise wording of nocebo verbal suggestions. The high heterogeneity in our sample indicates that unaccounted variables impacted the association between suggestibility and nocebo responding. Our inclusion criteria resulted in a diverse sample of outcomes (e.g., pain, motor control) and nocebo induction methods (e.g., verbal suggestion, conditioning), which might have contributed to the heterogeneity and limit the generalizability of our findings. Finally, most of the studies (*k*=6; *n* range: 46-93) were only sufficiently powered to detect moderate-to-large effects, and thus were insufficiently powered to reliably detect weak effects. This limitation may have also led to an underestimation of the association between suggestibility and nocebo responding. Insofar as the observed effect size is small in magnitude, this indicates that suggestibility is only one of multiple factors contributing to variability in response to nocebos including state anxiety (Rooney et al., 2022) and empathy (Meeuwis et al., 2023). It will be important in the future to evaluate whether these reflect independent or interacting effects. Cumulatively, these limitations highlight the need for rigorously designed experiments with meticulous control comparisons to further examine the apparent link between suggestibility and nocebo responses.

### Summary

This meta-analysis found a reliable association between trait suggestibility and nocebo responding. Given the small effect size, the present results raise important questions about the practical significance of this effect. However, owing to the relatively small number of studies, methodological heterogeneity, and inconsistent reporting of verbal suggestions, future research is needed to better assess the practical significance of suggestibility as a moderator of nocebo responding, particularly outside of an experimental context. From a clinical standpoint, understanding the role of suggestibility in nocebo responding and symptom perception more generally can pave the way for more personalized patient care. Highly suggestible individuals might be more susceptible to negative treatment information and thus could benefit from specific communication strategies that minimize nocebo effects (Arrow et al., 2022; Kari et al., 2021; Spiegel, 1997; Wells & Kaptchuk, 2012).

Integrating suggestibility into existing models of nocebo responding, as has been done in accounts of related conditions such as functional neurological disorder (Fiorio et al., 2022), could offer a more holistic understanding of variability in nocebo responses. By doing so, we can potentially uncover nuanced interactions between suggestibility and other established factors, such as the manner in which side effect information is communicated to participants and patients, as well as their level of trust Figin health care professionals. As we aim for more personalized and effective patient care and move towards an era of precision medicine (Seyhan & Carini, 2019), understanding individual differences factors that moderate nocebo responding is paramount for improving treatment outcomes.

## Data availability statement

All data relevant to the present meta-analysis is provided in the article. Data are freely available from previous research studies.

## References

1. Acunzo, D. J., & Terhune, D. B. (2021). A critical review of standardized measures of hypnotic suggestibility. Int J Clin Exp Hypn, 69(1), 50–71. 10.1080/00207144.2021.1833209

2. Arrow, K., Burgoyne, L. L., & Cyna, A. M. (2022). Implications of nocebo in anaesthesia care. Anaesthesia, 77 *Suppl 1*, 11–20. 10.1111/anae.15601

3. Bagaric, B., Jokic-Begic, N., & Sangster Jokic, C. (2022). The nocebo effect: A review of contemporary experimental research. Int J Behav Med, 29(3), 255–265. 10.1007/s12529-021-10016-y

4. Barber, T. X., & Wilson, S. C. (1978). The barber suggestibility scale and the creative imagination scale: Experimental and clinical applications. Am J Clin Hypn, 21(2-3), 84–108. 10.1080/00029157.1978.10403966

5. Bell, V., Oakley, D. A., Halligan, P. W., & Deeley, Q. (2011). Dissociation in hysteria and hypnosis: Evidence from cognitive neuroscience. J Neurol Neurosurg Psychiatry, 82(3), 332–339. 10.1136/jnnp.2009.199158

6. Bender, F. L., Rief, W., & Wilhelm, M. (2023). Really just a little prick? A meta-analysis on adverse events in placebo control groups of seasonal influenza vaccination rcts. Vaccine, 41(2), 294–303. 10.1016/j.vaccine.2022.11.033

7. Brascher, A. K., Sutterlin, S., Scheuren, R., Van den Bergh, O., & Witthoft, M. (2020). Somatic symptom perception from a predictive processing perspective: An empirical test using the thermal grill illusion. Psychosom Med, 82(7), 708–714. 10.1097/PSY.0000000000000824

8. Brass, M., Liefooghe, B., Braem, S., & De Houwer, J. (2017). Following new task instructions: Evidence for a dissociation between knowing and doing. Neuroscience & Biobehavioral Reviews, 81, 16–28. 10.1016/j.neubiorev.2017.02.012

9. Brooke, B. S., Schwartz, T. A., & Pawlik, T. M. (2021). Moose reporting guidelines for meta-analyses of observational studies. JAMA Surgery, 156(8), 787–788. 10.1001/jamasurg.2021.0522

10. Colloca, L., & Barsky, A. J. (2020). Placebo and nocebo effects. New England Journal of Medicine, 382(6), 554–561. 10.1056/NEJMra1907805

11. Colloca, L., & Miller, F. G. (2011). Harnessing the placebo effect: The need for translational research. Philos Trans R Soc Lond B Biol Sci, 366(1572), 1922–1930. 10.1098/rstb.2010.0399

12. Corsi, N., & Colloca, L. (2017). Placebo and nocebo effects: The advantage of measuring expectations and psychological factors. Front Psychol, 8, 308. 10.3389/fpsyg.2017.00308

13. Duval, S., & Tweedie, R. (2000). Trim and fill: A simple funnel-plot-based method of testing and adjusting for publication bias in meta-analysis. Biometrics, 56(2), 455–463. 10.1111/j.0006-341x.2000.00455.x

14. Egger, M., Davey Smith, G., Schneider, M., & Minder, C. (1997). Bias in meta-analysis detected by a simple, graphical test. BMJ, 315(7109), 629–634. 10.1136/bmj.315.7109.629

15. Fiorio, M., Braga, M., Marotta, A., Villa-Sanchez, B., Edwards, M. J., Tinazzi, M., & Barbiani, D. (2022). Functional neurological disorder and placebo and nocebo effects: Shared mechanisms. Nat Rev Neurol, 18(10), 624–635. 10.1038/s41582-022-00711-z

16. Haas, J. W., Bender, F. L., Ballou, S., Kelley, J. M., Wilhelm, M., Miller, F. G., Rief, W., & Kaptchuk, T. J. (2022). Frequency of adverse events in the placebo arms of covid-19 vaccine trials: A systematic review and meta-analysis. JAMA Network Open, 5(1), e2143955–e2143955. 10.1001/jamanetworkopen.2021.43955

17. Hauser, W., Hansen, E., & Enck, P. (2012). Nocebo phenomena in medicine: Their relevance in everyday clinical practice. Dtsch Arztebl Int, 109(26), 459–465. 10.3238/arztebl.2012.0459

18. Heller, M. K., Chapman, S. C. E., & Horne, R. (2022). Beliefs about medicines predict side-effects of placebo modafinil. Ann Behav Med, 56(10), 989–1001. 10.1093/abm/kaab112

19. Hilgard, E. R., Crawford, H. J., & Wert, A. (1979). The stanford hypnotic arm levitation induction and test (shalit): A six-minute hypnotic induction and measurement scale. International Journal of Clinical and Experimental Hypnosis, 27(2), 111–124. 10.1080/00207147908407551

20. Hohwy, J. (2020). New directions in predictive processing. Mind & Language, 35(2), 209–223. 10.1111/mila.12281

21. Jensen, M. P., Ehde, D. M., Hakimian, S., Pettet, M. W., Day, M. A., & Ciol, M. A. (2023). Who benefits the most from different psychological chronic pain treatments? An exploratory analysis of treatment moderators. J Pain, 24(11), 2024–2039. 10.1016/j.jpain.2023.06.011

22. Kari, A. L., Lauren, C. H., & Alia, J. C. (2021). Changing mindsets about side effects. BMJ Open, 11(2), e040134. 10.1136/bmjopen-2020-040134

23. Kern, A., Kramm, C., Witt, C. M., & Barth, J. (2020). The influence of personality traits on the placebo/nocebo response: A systematic review. Journal of Psychosomatic Research, 128, 109866. 10.1016/j.jpsychores.2019.109866

24. Khan, A. Y., Baade, L., Ablah, E., McNerney, V., Golewale, M. H., & Liow, K. (2009). Can hypnosis differentiate epileptic from nonepileptic events in the video/eeg monitoring unit? Data from a pilot study. Epilepsy Behav, 15(3), 314–317. 10.1016/j.yebeh.2009.04.004

25. Kotov, R. I., Bellman, S. B., & Watson, D. B. (2004). Multidimensional iowa suggestibility scale (miss) brief manual *Unpublished manuscript*.

26. Leigh, R., MacQueen, G., Tougas, G., Hargreave, F. E., & Bienenstock, J. (2003). Change in forced expiratory volume in 1 second after sham bronchoconstrictor in suggestible but not suggestion-resistant asthmatic subjects: A pilot study. Psychosom Med, 65(5), 791–795. 10.1097/01.psy.0000079454.48714.1b

27. Meeuwis, S. H., Wasylewski, M. T., Bajcar, E. A., Bieniek, H., Adamczyk, W. M., Honcharova, S., Di Nardo, M., Mazzoni, G., & Babel, P. (2023). Learning pain from others: A systematic review and meta-analysis of studies on placebo hypoalgesia and nocebo hyperalgesia induced by observational learning. Pain. 10.1097/j.pain.0000000000002943

28. Milling, L. S., Valentine, K. E., LoStimolo, L. M., Nett, A. M., & McCarley, H. S. (2021). Hypnosis and the alleviation of clinical pain: A comprehensive meta-analysis. International Journal of Clinical and Experimental Hypnosis. 10.1080/00207144.2021.1920330

29. Nasiri-Dehsorkhi, H., Vaziri, S., Esmaillzadeh, A., & Adibi, P. (2023). Psychological distress, perceived stress and nocebo effect (multifood adverse reaction) in irritable bowel syndrome patients. J Educ Health Promot, 12, 257. 10.4103/jehp.jehp_221_23

30. Nitzan, U., Chalamish, Y., Krieger, I., Erez, H. B., Braw, Y., & Lichtenberg, P. (2015). Suggestibility as a predictor of response to antidepressants: A preliminary prospective trial. J Affect Disord, 185, 8–11. 10.1016/j.jad.2015.06.028

31. Oakley, D. A., Walsh, E., Mehta, M. A., Halligan, P. W., & Deeley, Q. (2021). Direct verbal suggestibility: Measurement and significance. Conscious Cogn, 89, 103036. 10.1016/j.concog.2020.103036

32. Page, M. J., Moher, D., Bossuyt, P. M., Boutron, I., Hoffmann, T. C., Mulrow, C. D., Shamseer, L., Tetzlaff, J. M., Akl, E. A., Brennan, S. E., Chou, R., Glanville, J., Grimshaw, J. M., Hróbjartsson, A., Lalu, M. M., Li, T., Loder, E. W., Mayo-Wilson, E., McDonald, S., . . . McKenzie, J. E. (2021). Prisma 2020 explanation and elaboration: Updated guidance and exemplars for reporting systematic reviews. BMJ, 372, n160. 10.1136/bmj.n160

33. Parsons, R. D., Bergmann, S., Wiech, K., & Terhune, D. B. (2021). Direct verbal suggestibility as a predictor of placebo hypoalgesia responsiveness. Psychosom Med, 83(9), 1041–1049. 10.1097/psy.0000000000000977

34. Petersen, G. L., Finnerup, N. B., Colloca, L., Amanzio, M., Price, D. D., Jensen, T. S., & Vase, L. (2014). The magnitude of nocebo effects in pain: A meta-analysis. Pain, 155(8), 1426–1434. 10.1016/j.pain.2014.04.016

35. Petrovic, P. (2008). Placebo analgesia and nocebo hyperalgesia--two sides of the same coin? Pain, 136(1-2), 5–6. 10.1016/j.pain.2008.03.004

36. Polczyk, R. (2016). Factor structure of suggestibility revisited: New evidence for direct and indirect suggestibility. Current Issues in Personality Psychology, 4(2), 87–96. 10.5114/cipp.2016.60249

37. Riegel, B., Tönnies, S., Hansen, E., Zech, N., Eck, S., Batra, A., & Peter, B. (2021). German norms of the harvard group scale of hypnotic susceptibility, form a (hgshs:A) and proposal of a 5-item short-version (hgshs-5:G). International Journal of Clinical and Experimental Hypnosis, 69(1), 112–123. 10.1080/00207144.2021.1836645

38. Rooney, T., Sharpe, L., Todd, J., Richmond, B., & Colagiuri, B. (2022). The relationship between expectancy, anxiety, and the nocebo effect: A systematic review and meta-analysis with recommendations for future research. Health Psychol Rev, 1-28. 10.1080/17437199.2022.2125894

39. Rooney, T., Sharpe, L., Todd, J., Tang, B., & Colagiuri, B. (2023). The nocebo effect across health outcomes: A systematic review and meta-analysis. Health Psychol. 10.1037/hea0001326

40. Sapkota, R. P., Brunet, A., & Kirmayer, L. J. (2020). Characteristics of adolescents affected by mass psychogenic illness outbreaks in schools in nepal: A case-control study. Front Psychiatry, 11, 493094. 10.3389/fpsyt.2020.493094

41. Seyhan, A. A., & Carini, C. (2019). Are innovation and new technologies in precision medicine paving a new era in patients centric care? J Transl Med, 17(1), 114. 10.1186/s12967-019-1864-9

42. Sharav, Y., Haviv, Y., & Tal, M. (2023). Placebo or nocebo interventions as affected by hypnotic susceptibility. Applied Sciences, 13(2), 931. https://www.mdpi.com/2076-3417/13/2/931

43. Shor, R. E., & Orne, E. C. (1963). Norms on the harvard group scale of hypnotic susceptibility, form a. International Journal of Clinical and Experimental Hypnosis, 11(1), 39–47. 10.1080/00207146308409226

44. Spiegel, H. (1997). Nocebo: The power of suggestibility. Prev Med, *26*(5 Pt 1), 616-621. 10.1006/pmed.1997.0229

45. Stein, M. V., Holt, R., Wieder, L., & Terhune, D. B. (2023). Responsiveness to direct verbal suggestions and dissociation independently predict symptoms associated with environmental factors. Psychopathology, 56(4), 324–328.

46. Szigeti, B., Weiss, B., Rosas, F. E., Erritzoe, D., Nutt, D., & Carhart-Harris, R. (2024). Assessing expectancy and suggestibility in a trial of escitalopram v. Psilocybin for depression. Psychol Med, 1-8. 10.1017/s0033291723003653

47. Terhune, D. B., Cleeremans, A., Raz, A., & Lynn, S. J. (2017). Hypnosis and top-down regulation of consciousness. Neuroscience and Biobehavioral Reviews, 81(Part A), 59–74. 10.1016/j.neubiorev.2017.02.002

48. The jamovi project. (2023). *Jamovi (version 2.3) [computer software]. Retrieved from* https://www.Jamovi.Org.

49. Thompson, T., Terhune, D. B., Oram, C., Sharangparni, J., Rouf, R., Solmi, M., Veronese, N., & Stubbs, B. (2019). The effectiveness of hypnosis for pain relief: A systematic review and meta-analysis of 85 controlled experimental trials. Neuroscience and Biobehavioral Reviews, 99, 298–310. 10.1016/j.neubiorev.2019.02.013

50. Van den Bergh, O., Witthoft, M., Petersen, S., & Brown, R. J. (2017). Symptoms and the body: Taking the inferential leap. Neurosci Biobehav Rev, 74(Pt A), 185–203. 10.1016/j.neubiorev.2017.01.015

51. Webster, R. K., Weinman, J., & Rubin, G. J. (2016). A systematic review of factors that contribute to nocebo effects. Health Psychol, 35(12), 1334–1355. 10.1037/hea0000416

52. Wells, R. E., & Kaptchuk, T. J. (2012). To tell the truth, the whole truth, may do patients harm: The problem of the nocebo effect for informed consent. Am J Bioeth, 12(3), 22–29. 10.1080/15265161.2011.652798

53. Wickramasekera, I. E., & Szlyk, J. P. (2003). Could empathy be a predictor of hypnotic ability? International Journal of Clinical and Experimental Hypnosis, 51(4), 390–399. 10.1076/iceh.51.4.390.16413

54. Wieder, L., Brown, R., Thompson, T., & Terhune, D. B. (2021). Suggestibility in functional neurological disorder: A meta-analysis. *Journal of Neurology*, Neurosurgery and Psychiatry, 92(2), 150–157. 10.1136/jnnp-2020-323706

55. Winter, D., & Braw, Y. (2022). Covid-19: Impact of diagnosis threat and suggestibility on subjective cognitive complaints. Int J Clin Health Psychol, 22(1), 100253. 10.1016/j.ijchp.2021.100253

56. Wolters, F., Peerdeman, K. J., & Evers, A. W. M. (2019). Placebo and nocebo effects across symptoms: From pain to fatigue, dyspnea, nausea, and itch. Front Psychiatry, 10, 470. 10.3389/fpsyt.2019.00470

57. Woody, E. Z., & Barnier, A. J. (2008). Hypnosis scales for the twenty-first century: What do we need and how should we use them? In The oxford handbook of hypnosis: Theory, research, and practice. (pp. 255-280). Oxford University Press.

58. Woody, E. Z., Drugovic, M., & Oakman, J. M. (1997). A reexamination of the role of nonhypnotic suggestibility in hypnotic responding. Journal of Personality and Social Psychology, 72(2), 399–407. 10.1037/0022-3514.72.2.399

59. Zech, N., Scharl, L., Seemann, M., Pfeifer, M., & Hansen, E. (2022). Nocebo effects of clinical communication and placebo effects of positive suggestions on respiratory muscle strength. Front Psychol, 13, 825839. 10.3389/fpsyg.2022.825839

60. Zech, N., Schrödinger, M., Seemann, M., Zeman, F., Seyfried, T. F., & Hansen, E. (2020). Time-dependent negative effects of verbal and non-verbal suggestions in surgical patients-a study on arm muscle strength. Front Psychol, 11, 1693. 10.3389/fpsyg.2020.01693

61. Zech, N., Seemann, M., Grzesiek, M., Breu, A., Seyfried, T. F., & Hansen, E. (2019). Nocebo effects on muscular performance - an experimental study about clinical situations. Front Pharmacol, 10, 219. 10.3389/fphar.2019.00219

